# The effect of trauma quality improvement programme implementation on quality of life among trauma patients in urban India

**DOI:** 10.1101/2024.09.28.24314529

**Authors:** Eelika Kapitan, Johanna Berg, Siddarth David, L N Mohan, Li Felländer-Tsai, Shamita Chatterjee, Ulf Ekelund, Nobhojit Roy, Max Petzhold, Johan Von Schreeb, Kapil Dev Soni, Sendhil Rajan, Monty Khajanchi, Martin Gerdin Wärnberg

## Abstract

Trauma accounts for around 4.4 million deaths annually and is among the leading causes of death, disproportionately affecting low- and middle-income countries. Trauma quality improvement programmes may reduce adverse patient outcomes but lack robust evidence. This study assessed the effect of implementing a trauma quality improvement programme using audit filters on health-related quality of life in adult trauma patients in urban India.

We conducted a controlled interrupted time series study (ClinicalTrials.gov ID NCT03235388) across four tertiary care university hospitals. Adult (≥18 years) trauma patients, who were alive at 90 days post-admission, were included. After a 14-month observation phase, two hospitals were randomized to the intervention arm and implemented the trauma quality improvement programme, while two control hospitals did not implement any interventions. The study spanned from 2017 to 2022.

Health-related quality of life was assessed 90 days post-admission using the EuroQol 5-dimensions (EQ-5D) questionnaire. The main outcome was EQ-5D health status, scaled from 0 to 100, analyzed using a difference-in-differences (DiD) approach. Descriptive analysis was performed for the five individual EQ-5D dimensions.

2307 participants were eligible for the analyses. The DiD analysis showed a statistically significant reduction in the EQ-5D health status score in the intervention arm (DiD estimate −4.07 [95% CI: −7.00; −1.15]). For the 819 patients with major trauma, there was a more pronounced, but statistically insignificant reduction (DiD estimate −4.86 [95% CI: −9.87; 0.15]). In the intervention arm, mobility, self-care, and pain/discomfort improved, while usual activities and anxiety/depression worsened.

Implementing trauma quality improvement programmes using audit filters may reduce the average health-related quality of life of trauma patients if more severely injured patients survive, highlighting the importance of assessing quality of life after the implementation of such programmes and ensuring adequate rehabilitation resources to support long-term recovery.

## Introduction

Trauma, defined as physical injury and the body’s subsequent response to it, accounts for 10% of disability-adjusted life-years (1). In general, the burden of injury declines as socio-demographic index increases, but certain types of injuries such as road injury, interpersonal violence and self-harm deviate from this trend, indicating the presence of other underlying factors that influence injury outcomes (1).

Disability following trauma can impact quality of life (QoL), return to work, and social participation, and the QoL for trauma patients often fails to return to pre-injury levels (2,3). Temporary or permanent disabilities impose an economic burden and hidden costs on patients and their families, predominantly due to out-of-pocket expenses for medicines and diagnostic tests, as well as financial strain from missed workdays of caregivers, often forcing patients to prioritize certain treatments over others and further affecting their QoL (4–6).

In addition to prevention, trauma care is important to reduce mortality and morbidity (7). A systematic review and meta-analysis found that establishing trauma quality improvement programmes may reduce mortality in trauma patients, although the evidence supporting this conclusion was of low quality since most included studies were observational, and no measures on morbidity were reported (8).

The World Health Organization (WHO) recommends implementing audit filters, pre-established variables that define deviations from the standard of care and may identify potential problems, as part of trauma quality improvement programmes across the world (9). However, high-level evidence supporting the effectiveness of quality improvement programs using audit filters on patient outcomes is lacking (10), and implementing these filters is time- and resource-intensive, with their usefulness being questioned (11,12).

To address this gap, we conducted the Trauma Audit Filters Trial (TAFT), a controlled interrupted time series study, across four tertiary hospitals in urban India to evaluate the effect of implementing a trauma quality improvement programme using audit filters on various patient outcomes (13). Following the intervention, there was an absolute reduction of 14% in the 30-day mortality rate among all patients, and a 31% reduction among patients with major trauma (13).

In addition to mortality, we also studied QoL as one of the pre-specified secondary outcomes of TAFT, so in this study, our aim was to assess the effect of implementing a trauma quality improvement programme using audit filters on health-related QoL among trauma patients.

## Methods

### Study design and setting

We did a secondary analysis of the TAFT (ClinicalTrials.gov ID NCT03235388), a controlled interrupted time series study conducted across four tertiary hospitals in urban India from October 2017 to October 2022. Each hospital had around 1500 beds and the clinical specialties relevant to trauma care available on-site. None of the hospitals were using any structured trauma quality improvement programmes upon enrollment. The primary outcome was all-cause mortality within 30-days of arrival to participating hospital, reported in a separate publication (13), and QoL at 90 days after arrival was one of the secondary outcomes. For reporting, this article follows the TREND (Transparent Reporting of Evaluations with Nonrandomized Designs) statement (14).

TAFT included three distinct phases. During the observation phase (months 1-14), a baseline was established for primary and secondary outcomes. During the implementation phase (months 15-20), hospitals were randomized to control and intervention arms in pairs - the hospital with the highest patient volume was paired with the hospital with the lowest patient volume and the two remaining hospitals were assigned to the second pair. During the intervention phase (months 21-42), the elected audit filter review board conducted monthly review meetings in the intervention hospitals, while control hospitals remained unchanged from the observation and implementation phase.

### Participants

For inclusion in TAFT, participants had to be adults (≥18 years) admitted with a history of trauma, defined as having any external causes of mortality and morbidity as per International Classification of Disease version 10 (ICD-10) codebook chapter 20 block V01-Y36. To assess the effect of audit filters on QoL, we included only participants who consented to follow-up and who had available information about surviving until 90 days post-admission.

### Intervention

The intervention involved developing and implementing trauma audit filters, reported elsewhere (13,15). In brief, the two hospitals randomized to the intervention arm participated in a two-day course to introduce the relevance and rationale for using trauma quality improvement programmes, including audit filters, and participated in an anonymous online Delphi survey to develop context specific audit filters (15). As a potential confounder that was not known when the study commenced, a new trauma center was established in one of the intervention arm hospitals with 168 beds for the trauma patients, including 24 intensive care unit beds, nine months after the start of intervention.

### Outcomes

The outcome was health-related QoL at 90 days after admission, recorded using the instrument EuroQol 5-dimensions 3-levels (EQ-5D-3L). This instrument includes a Visual Analogue Scale (EQ-VAS) to measure overall health status, and a descriptive system (16). The EQ-VAS, the main outcome of this study, ranges from 0 to 100, with 0 representing the worst imaginable health, and 100 the best (17). The descriptive system of EQ-5D consists of five dimensions - mobility, self-care, usual activities, pain or discomfort, and anxiety or depression (16). Each of these five dimensions are rated according to the same three-level likert scale: no problems, some problems, and extreme problems (16). The five dimensions were considered secondary outcomes of this study. The EQ-5D-3L instrument was chosen over the EQ-5D-5L because it is shorter, making data collection more feasible.

### Covariates

To account for possible differences in sample characteristics between the hospitals and before and after the intervention within the study arm, the following covariates were included in the descriptive statistics: age, sex, major trauma (defined as those admitted for over three days with an injury severity score [ISS] >15), type of injury, mechanism of injury, Glasgow Coma Scale (GCS), ISS, and days in hospital. Age, sex, type of injury, GCS, and ISS were also considered potential confounders in the difference-in-differences analyses and were therefore adjusted for.

### Data collection and management

The data collection was conducted by the project officers stationed in the emergency departments of four hospitals. The project officers were not blinded to the intervention assignment. Information on covariates was collected either by direct observation or from the participant’s hospital record. Since TAFT utilized both prospective and retrospective data, only participants who were enrolled prospectively and had consented for follow-up were included in the QoL analyses. After 90-days post-admission, the project officers telephonically collected the QoL information, using the EQ-5D-3L instrument in the language spoken by the participants. To ensure participants’ cognitive abilities, the officers first introduced themselves, reminded participants about the study, and made general inquiries about their well-being before administering the questionnaire.

### Sample size

QoL at 90 days was collected as a secondary outcome within a larger study, thus we did not conduct any a priori sample size calculations.

### Statistical methods

Data analysis was conducted using R version 4.3.3 with the confidence level set at 95% and the significance level set at 5%. Pre- and post-intervention study population characteristics were reported across the control and intervention arm using descriptive statistics to detect potential differences in sample characteristics. Missing data across descriptive variables for the control and intervention arm were tabulated. Participants with any missing data in any of the six outcome variables were considered as incomplete cases and excluded from the analyses, which allowed us to analyze the same cohort for all outcome variables.

We employed a quasi-experimental difference-in-differences approach that is suited for evaluating policy changes or interventions as it accounts for both time trends and unobserved confounders (18). This approach relies on the assumption of common outcome trends between study arms in the absence of the intervention, enabling us to isolate the effect of the intervention even when confounders varying across study arms are time-invariant, and time-varying confounders are study arm invariant (19). Data from the intervention and control arm hospitals were pooled separately. To compare changes in EQ-5D health status scores between the study arms, we used a generalized linear model as follows:

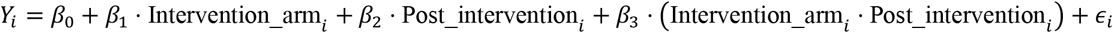

where *β*_0_ is the baseline average for the control arm before intervention, *β*_1_ is the difference between the intervention and control arm before intervention, β_2_ is the time trend in control arm, *β*_3_ is the difference in EQ-5D health status changes over time between intervention and control arm, and ϵ_i_ is the error term. Both unadjusted and adjusted models (for age, sex, type of injury, GCS, and ISS) were included in the analysis.

Additionally, the proportions of participants with some problems and extreme problems in each of the five dimensions were reported before and after the intervention.

### Subgroup analyses

Given the particularly significant reduction in mortality among patients with major trauma after the intervention (13), we conducted a subgroup analysis focusing on patients with major trauma only.

### Ethical Considerations

TAFT was approved by all participating hospitals in India: Maulana Azad Medical College (MAMC) - approved 2017/07/19, F.1/IEC/MAMC/(57/02/2017/No 113. SSKM/IPGME&R, Kolkata - approved 2017/08/21, IPGME&R/IEC/2017/396. JJ Hospital, Mumbai - approved 2017/08/22, No. IEC/Pharm/CT/111/A/2017. St John’s Medical College, Bangalore - approved 2017/08/24, Ref. no. 160/2017, and the Swedish Ethical Review Authority - approved 2017/06/07, 2017/930-31/2.

## Results

### Participants

A total of 10 143 study participants were included in TAFT between October 2017 and October 2022. After including only those whose data were collected prospectively, who consented to follow-up and who had available information about surviving until 90 days post-admission and excluding those with missing data on any of the six outcome variables, the final study population consisted of 2307 participants (Figure 1).

**Figure 1.**
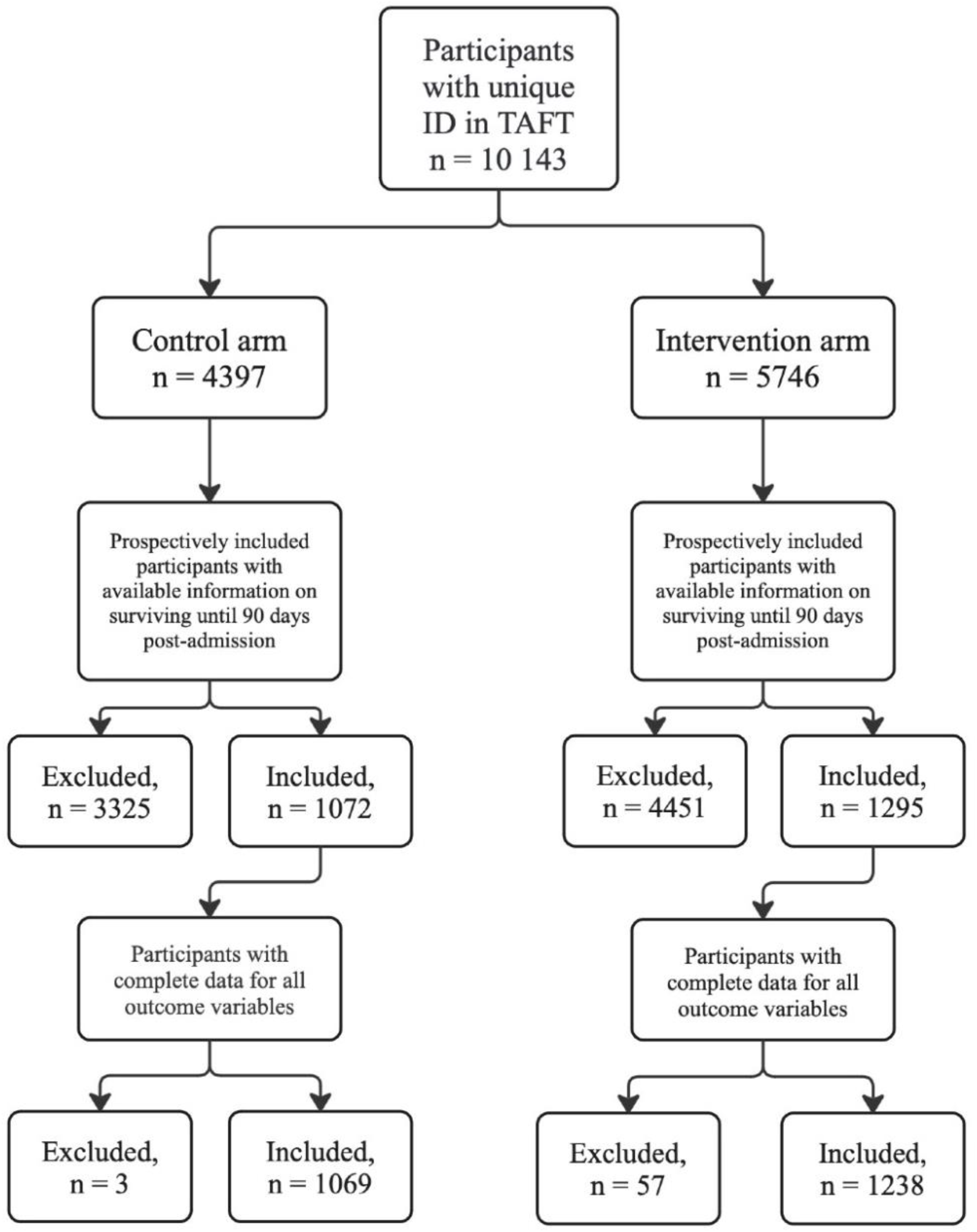
Flow diagram of the study population by study arm

### Descriptive data

Out of the total 2307 participants in the study population, 1069 participants were in the control arm and 1238 participants were in the intervention arm (Table 1). The participants in the two study arms were comparable in sex distribution, type of injury, GCS and length of hospital stay. Participants in the intervention arm compared to the control arm were older and had more often major trauma with higher ISS. The most prominent differences in sample characteristics were seen between the pre-intervention and post-intervention group in the intervention arm, where the proportion of participants with major trauma increased from 43% in the pre-intervention group to 72% in the post-intervention group, transport accident cases increased from 55% to 80%, the mean GCS decreased by 1.5 points, the mean ISS increased by 2.7 points, and the mean number of days spent in hospital increased by 1.4 days in the pre- and post-intervention groups. The mean EQ-5D health status increased by 1.3 points in the control arm and decreased by 2.8 points in the intervention arm.

**Table 1.** Sample characteristics across study arms, before and after the intervention (n = 2307)

| **Variable** | **Control Arm** |  |  | **Intervention Arm** |  |  |
| --- | --- | --- | --- | --- | --- | --- |
|  | **Overall, n = 1,069<sup>a</sup>** | **Before, n = 446<sup>a</sup>** | **After, n = 623<sup>a</sup>** | **Overall, n = 1,238<sup>a</sup>** | **Before, n = 380<sup>a</sup>** | **After, n = 858<sup>a</sup>** |
| **Age** | 36.3 (15.4) | 35.7 (15.4) | 36.7 (15.4) | 38.4 (14.4) | 38.9 (15.5) | 38.2 (13.9) |
| Missing | 1 | 0 | 1 |  |  |  |
| **Sex** |  |  |  |  |  |  |
| Female | 193 (18%) | 77 (17%) | 116 (19%) | 223 (18%) | 98 (26%) | 125 (15%) |
| Male | 876 (82%) | 369 (83%) | 507 (81%) | 1,015 (82%) | 282 (74%) | 733 (85%) |
| **Major trauma** | 151 (16%) | 69 (17%) | 82 (15%) | 668 (64%) | 114 (43%) | 554 (72%) |
| Missing | 108 | 31 | 77 | 196 | 112 | 84 |
| **Type of injury** |  |  |  |  |  |  |
| Blunt | 1,035 (97%) | 425 (95%) | 610 (98%) | 1,226 (99%) | 373 (98%) | 853 (99%) |
| Penetrating | 29 (2.7%) | 17 (3.8%) | 12 (1.9%) | 12 (1.0%) | 7 (1.8%) | 5 (0.6%) |
| Both blunt and penetrating | <5 | <5 | 0 (0%) | 0 (0%) | 0 (0%) | 0 (0%) |
| Missing | 1 | 0 | 1 |  |  |  |
| **Mechanism of injury** |  |  |  |  |  |  |
| Assault | 70 (6.5%) | 32 (7.2%) | 38 (6.1%) | 63 (5.1%) | 31 (8.2%) | 32 (3.7%) |
| Burns | 44 (4.1%) | 13 (2.9%) | 31 (5.0%) | 16 (1.3%) | 9 (2.4%) | 7 (0.8%) |
| Fall | 191 (18%) | 78 (17%) | 113 (18%) | 230 (19%) | 105 (28%) | 125 (15%) |
| Intentional self-harm | 5 (0.5%) | <5 | <5 | <5 | <5 | <5 |
| Other accident, not violence | 167 (16%) | 92 (21%) | 75 (12%) | 25 (2.0%) | 21 (5.5%) | <5 |
| Transport accident | 587 (55%) | 229 (51%) | 358 (57%) | 899 (73%) | 210 (55%) | 689 (80%) |
| Undetermined intent | <5 | 0 (0%) | <5 | <5 | <5 | 0 (0%) |
| Missing | 2 | 0 | 2 | 2 | 2 | 0 |
| **Glasgow Coma Scale** | 14.7 (1.6) | 14.7 (1.6) | 14.6 (1.7) | 13.3 (2.2) | 14.3 (1.8) | 12.8 (2.1) |
| Missing | 20 | 9 | 11 | 1 | 1 | 0 |
| **Injury Severity Score (ISS)** | 6.7 (7.9) | 6.6 (8.1) | 6.7 (7.7) | 11.9 (7.3) | 10.0 (8.2) | 12.7 (6.8) |
| Missing | 84 | 24 | 60 | 157 | 82 | 75 |
| **Days in hospital** | 10.9 (14.4) | 10.9 (11.5) | 10.9 (16.1) | 8.4 (13.3) | 7.4 (21.4) | 8.8 (7.2) |
| Missing | 28 | 8 | 20 | 7 | 2 | 5 |
| **EQ5D Health Status at 90 days<sup>b</sup>** | 80.8 (19.5) | 80.0 (20.1) | 81.3 (19.1) | 77.8 (14.7) | 79.7 (18.2) | 76.9 (12.8) |

<sup>a</sup>Mean (SD); n (%)<sup>b</sup>Scale ranges from 0 to 100, with 0 being the worst imaginable health, and 100 the best.

### Difference-in-differences of EQ-5D health status score

Both the unadjusted and adjusted difference-in-differences analyses showed that the intervention was associated with a statistically significant reduction in the EQ-5D health status (Table 2). In the unadjusted analysis, the EQ-5D health status was reduced by 4.07 units (95% CI: −7.00; −1.15) and in the adjusted analysis, the EQ-5D health status was reduced by 3.41 units (95% CI: −6.54; −0.27). Among the patients with major trauma, the intervention was associated with more pronounced, but statistically insignificant reduction in the EQ-5D health status. In the unadjusted analysis, the EQ-5D health status was reduced by 4.86 units (95% CI: −9.87; 0.15), and by 4.52 units (95% CI: −9.77; 0.74) in the adjusted analysis.

**Table 2.** Difference-in-differences of EQ-5D health status score.

|  | **All patients**<br>**n = 2307** |  | **Major trauma patients<sup>a</sup>**<br>**n = 819** |  |
| --- | --- | --- | --- | --- |
| Variable | $\beta$(95% CI)<sup>b</sup> | p-value | $\beta$(95% CI)<sup>b</sup> | p-value |
| **UNADJUSTED** |  |  |  |  |
| Intervention effect | -4.07 (-7.00; -1.15) | **0.006** | -4.86 (-9.87; 0.15) | 0.06 |
| **ADJUSTED** |  |  |  |  |
| Intervention effect | -3.41 (-6.54; -0.27) | **0.03** | -4.52 (-9.77; 0.74) | 0.09 |
| Age | -0.17 (-0.22; -0.12) | **<0.001** | -0.07 (-0.14; -0.01) | **0.03** |
| Sex |  |  |  |  |
| *Female* | NA | NA | NA | NA |
| *Male* | -0.29 (-2.20; 1.61) | 0.76 | -1.27 (-3.66; 1.12) | 0.3 |
| Type of injury |  |  |  |  |
| *Blunt* | NA | NA | NA | NA |
| *Penetrating* | 8.79 (2.76; 14.82) | **0.004** | 21.80 (6.62; 36.99) | **0.005** |
| *Both blunt and penetrating* | -0.88 (-17.22; 15.47) | 0.92 | 2.34 (-23.55; 28.23) | 0.86 |
| Glasgow Coma Scale | 0.28 (-0.10; 0.66) | 0.15 | 0.64 (0.21; 1.06) | **0.004** |
| Injury Severity Score | -0.15 (-0.25; -0.05) | **0.003** | -0.39 (-0.67; -0.11) | **0.006** |

<sup>a</sup>Major trauma patients – those admitted for over three days with an ISS >15<sup>b</sup>CI = Confidence Interval

### Descriptive analysis of the five EQ-5D dimensions

In the intervention arm, there was a decrease in experiencing some problems with mobility (from 33% to 30%), self-care (from 29% to 14%) and pain or discomfort (from 61% to 45%), but an increase in problems with usual activities (from 39% to 62%) and anxiety or depression (from 31% to 76%) (Figure 2). Further, there was a decrease in experiencing extreme problems in all dimensions in the intervention arm. The control arm had minor changes across the EQ-5D dimensions before and after the intervention.

**Figure 2.**
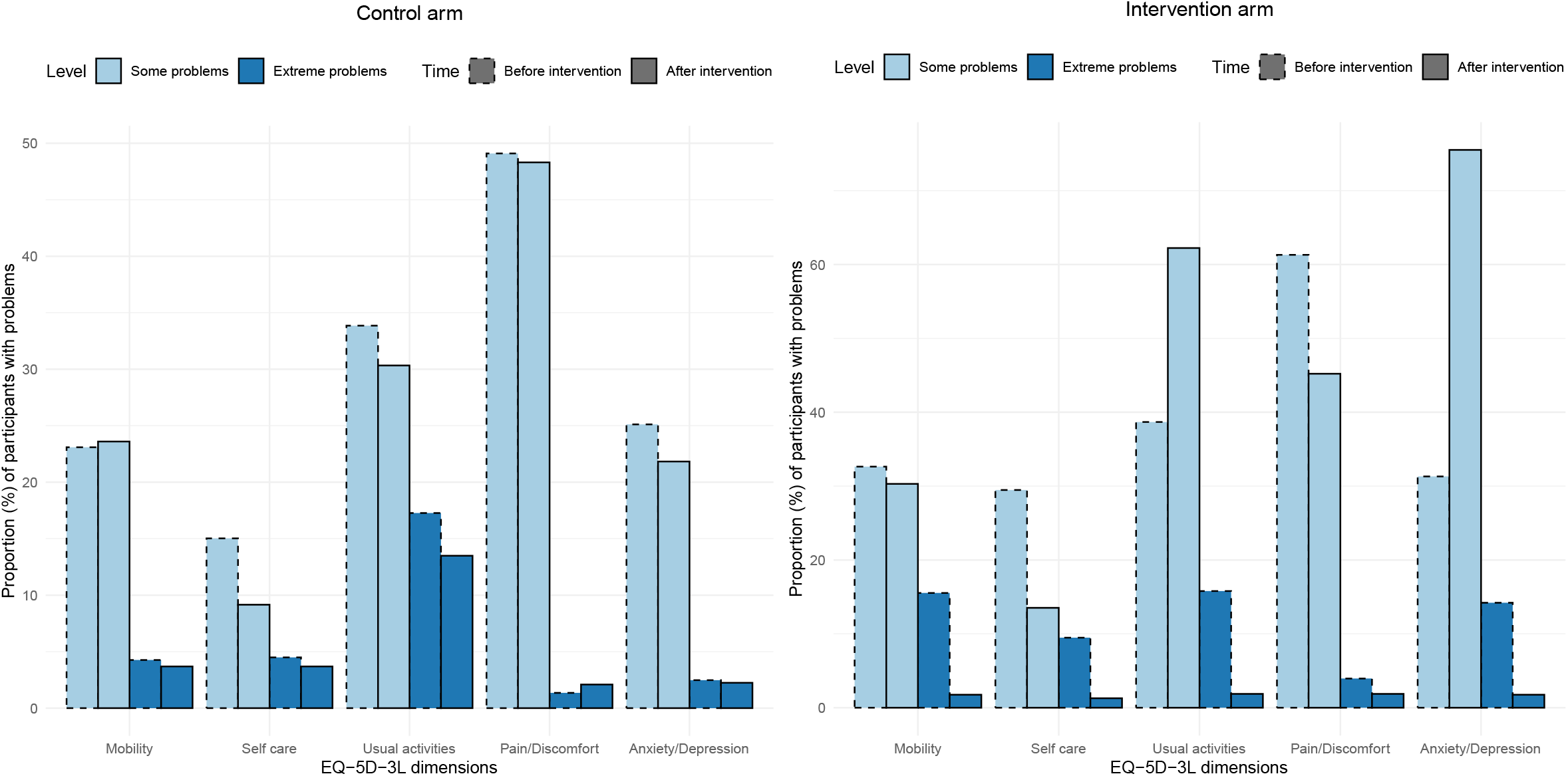
Proportions of participants with some and extreme problems in each EQ-5D dimension before and after the intervention (control arm n = 1069; intervention arm n = 1238)

## Discussion

We found that the overall EQ-5D health status declined after implementing a trauma quality improvement programme using audit filters. The effects on the individual EQ-5D dimensions were surprisingly heterogeneous with improvements in mobility, self-care, and pain but with increased anxiety or depression, and inability to perform usual activities. The reduced QoL health status may be attributable to survival bias. Improved survival was observed following the intervention, which was particularly pronounced in patients with severe trauma (13), likely being the key driver of the observed changes in QoL within the intervention arm. This pronounced effect on survival in patients with severe trauma could be mediated using audit filters, focusing primarily on early-stage trauma management such as resuscitation and airway management (15).

While improved survival of the severely injured is a positive outcome, it introduces a multitude of new challenges for patients, which may explain the reduced QoL scores. Improved survival can lead to extended needs of rehabilitative services, which is often inadequate in resource-poor countries (4,20). A qualitative study of trauma patients in urban India found poor public hospital conditions and barriers like transportation costs often deter follow-up care, resulting in a greater burden once patients return home (4). This may explain reduced QoL health scores observed in our study, particularly among the severely injured requiring more rehabilitation. Beyond extended rehabilitation, trauma survivors often encounter substantial socioeconomic and functional difficulties. Factors like lost wages, forced unemployment, and a lack of adequate insurance coverage contribute to financial toxicity, further exacerbating declines in QoL (21). With more severely injured patients surviving post-intervention (13), these financial challenges may have become more pronounced, reflecting the decline in health-related QoL. Additionally, surviving severe trauma can lead to psychosocial and role functioning difficulties and impairments in basic functions like feeding, bathing, and mobility issues (22,23), and patients who survived severe injuries post-intervention may have experienced these challenges more substantially.

The decline in QoL health status in our study may suggest that trauma patients, particularly patients with severe trauma, experienced delays in reaching recovery milestones after the intervention, possibly influencing the trajectory and timeliness of recovery. A Vietnamese study on QoL and recovery patterns among trauma patients identified four recovery periods: an acute treatment period at one month, a rehabilitation period at two months, an adaptation period at four months, and a stable end period at 12 months (24), and a similar pattern was identified in an Indian cohort study on mortality and QoL following injuries (3). Reduced mortality among the severely injured after the intervention (13) likely delayed their progression through these recovery stages compared to patients in the pre-intervention group. Many may not have advanced beyond the rehabilitation period by the 90-day follow-up, contributing to the lower overall health status scores observed.

The descriptive analysis of EQ-5D dimensions showed heterogeneous effects of the intervention. Improvements were observed in mobility, self-care, and pain or discomfort, but more participants reported having some problems with usual activities and anxiety or depression. The decline in performing usual activities, but improvement in mobility and pain may suggest that factors beyond physical capability may be involved. One aspect of usual activities is working (25) and given that the participants were mostly working-age males of lower socioeconomic status, returning to work after surviving a severe trauma may have been more challenging. This pattern aligns with findings from a qualitative study on the lived experiences of trauma patients in urban India, where returning to work was viewed as a key indicator of recovery, symbolizing both a return to pre-trauma normalcy and a crucial factor in economic stability (4). The inability or reduced capacity to work may have contributed to the economic burden experienced by trauma victims and their families, ultimately impacting mental health, and contributing to a decline in the anxiety or depression dimension, an association that has also been suggested earlier (21,26,27,28).

Trauma audit filters were employed to detect opportunities for improving trauma care by discussing flagged cases with the review board and making changes accordingly. While the changes in trauma care may extend beyond the trauma audit filters, these implemented audit filters did not address any aspects of the rehabilitation process that may have had a direct impact on health-related QoL (15). Future studies could explore the underlying causes of increased psychological morbidity and the decrease in performing usual activities among injured patients in contexts comparable to urban India, to better tailor rehabilitation services.

### Strengths and limitations

This is the first study that looks at the effect of quality improvement programmes using audit filters on the QoL, as previous research has predominantly focused on mortality outcomes (8). Additionally, one of the major strengths of this study is the use of controlled interrupted time series study design, which is among the strongest quasi-experimental study designs for evaluating quality improvement interventions in the healthcare system when a randomized controlled trial is not feasible (29).

Our study also has several limitations. First, the absence of data on survival status for some participants could introduce selection bias. If the post-intervention participants were surviving more severe injuries and were less willing to be followed up on, it might have led to an overestimation of the intervention effect. Second, the use of EQ-5D-3L instead of EQ-5D-5L might have led to a systematic overestimation of health problems due to higher cut-points and reduced refinement (30). Third, the lack of sociodemographic data limits our understanding and control of their influence on QoL outcomes, and factors such as education, employment, marital status, and pre-existing illnesses could be potential confounders (31). Fourth, the establishment of a new trauma center in the intervention arm could introduce significant confounding, as it is difficult to differentiate the effects between the increased capabilities of the trauma center and the quality improvement programme. Fifth, observer bias might have affected the results, as project officers aware of the intervention might have been more diligent in identifying health issues in the intervention arm. Finally, the 90-day follow-up period was relatively short. Previous research indicates that trauma impacts on QoL can extend beyond three months, often persisting far more than a year (4,32,33).

## Conclusion

Our findings indicate that overall QoL health status, as measured by EQ-5D-3L, declined following the implementation of a trauma quality improvement programme, possibly due to improved survival among severely injured patients which consequently extended rehabilitation needs. The effects on individual QoL dimensions were more varied, with improvements in mobility, self-care, and pain or discomfort, but declines in usual activities and anxiety or depression. These results underscore the significance of assessing health-related QoL of trauma patients after trauma quality improvement programme implementation to understand how to support them effectively and emphasize the growing need for expanded rehabilitation services to facilitate patients’ long-term recovery.

## Data Availability

All data produced in the present work are contained in the manuscript.

## Notes

### Competing Interest Statement

The authors have declared no competing interest.

### Clinical Trial

NCT03235388

### Funding Statement

This work was funded by the Swedish Research Council (2016-02041).

### Author Declarations

The Swedish Ethical Review Authority gave ethical approval for this work (2017/930-31/2). The local ethical review board of Maulana Azad Medical College (MAMC) gave ethical approval for this work (F.1/IEC/MAMC/(57/02/2017/No 113). The local ethical review board of Institute of Post-Graduate Medical Education and Research and Seth Sukhlal Karnani Memorial Hospital (IPGME&R/SSKM), Kolkata gave ethical approval for this work (IPGME&R/IEC/2017/396). The local ethical review board of JJ Hospital, Mumbai gave ethical approval for this work (No. IEC/Pharm/CT/111/A/2017). The local ethical review board of St Johns Medical College, Bangalore gave ethical approval for this work (Ref. no. 160/2017).

### Summary of Updates

One co-author was removed from the list of authors as they were invited to serve as a co-guest editor for a special issue of the journal where this paper will be submitted. This adjustment was made to avoid any potential conflict of interest.

